# Methicillin-resistant *Staphylococcus aureus* in Saudi Arabia: genomic evidence of recent clonal expansion and plasmid-driven resistance dissemination

**DOI:** 10.1101/2025.01.31.25321315

**Authors:** Ahmed Yousef Alhejaili, Ge Zhou, Heba Halawa, Jiayi Huang, Omniya Fallatah, Raneem Hirayban, Sara Iftikhar, Abrar AlAsmari, Mathew Milner, Manuel Banzhaf, Albandari A. Alzaidi, Ahmad A. Rajeh, Maram Abdulmohsen Al-Otaiby, Sarah S. Alabbad, Doua Bukhari, Abdullah N. Aljurayyan, Alanoud T. Aljasham, Zeyad A. Alzeyadi, Sulaiman M. Alajel, Rawan Hamdan Alanazi, Majed Alghoribi, Mashal M. Almutairi, Arnab Pain, Abiola Senok, Danesh Moradigaravand, Waleed Al Salem

## Abstract

**Objectives:** *Staphylococcus aureus* is a leading cause of hospital-acquired infections worldwide. Over recent decades, methicillin-resistant *Staphylococcus aureus* (MRSA), which is resistant to multiple antimicrobials, has emerged as a significant pathogenic strain in both hospital and community settings. The rapid emergence and dissemination of MRSA clones are driven by a dynamic and evolving population, spreading swiftly across regions on epidemiological time scales. Despite the vast geographical expanse and diverse demographics of the Kingdom of Saudi Arabia and the broader West Asia region, the population diversity of MRSA in hospitals in these areas remains underexplored.

**Methods:** We conducted a large-scale genomic analysis of a systematic *Staphylococcus aureus* collection obtained from 34 hospitals across all provinces of KSA, from diverse infection sites between 2022 and 2024. The dataset comprised 582 MRSA and 30 methicillin-susceptible *Staphylococcus aureus* (MSSA) isolates, all subjected to whole-genome sequencing. A combination of phylogenetic and population genomics approaches was utilized to analyze the genomic data. Hybrid sequencing approach was employed to retrive the complete plasmid content.

**Results:** The population displayed remarkable diversity, comprising 35 distinct sequence types (STs), with the majority harboring community-associated SCCmec loci (types IVa, V/VII, and VI). Virulence factors associated with community-acquired MRSA (CA-MRSA), including Panton-Valentine Leukocidin (PVL) genes, were identified in 12 distinct STs. Dominant clones, including ST8-t008 (USA300), ST88-t690, ST672-t3841, ST6-t304, and ST5-t311, were associated with infections at various body sites and were widely disseminated across the country. Linezolid and vancomycin resistance were mediated by *cfr*-carrying plasmids and mutations in the *vraR* gene (involved in cell-wall stress response) and the *murF* gene (peptidoglycan biosynthesis) in five isolates, respectively. Phylodynamic analysis revealed rapid expansion of the dominant clones, with their emergence estimated to have occurred 10–20 years ago. Plasmidome analysis uncovered a diverse repertoire of *blaZ*-containing plasmids and the sharing of *erm(C)*-encoding plasmids among major clades. The acquisition of plasmids coincided with clonal expansion.

**Conclusions:** Our results highlight the recent concurrent expansion and geographical dissemination of CA-MRSA clones across hospitals. These findings also underscore the interplay between clonal spread and horizontal gene transfer in shaping the resistance landscape of MRSA.

## Introduction

*Staphylococcus aureus* is a major human pathogen responsible for a wide spectrum of infections in hospital settings worldwide. Its clinical significance has been amplified by the emergence of pandemic clones, driven by the acquisition of resistance genes under the selective pressure of antimicrobial use in healthcare environments. Notably, in response to methicillin use over recent decades, methicillin-resistant S. aureus (MRSA) has emerged, characterized by the acquisition of the *mecA* gene within the staphylococcal cassette chromosome *mec* (SCC*mec*) [1]. Globally circulating MRSA clones impose a substantial burden on healthcare systems, contributing to increased mortality, prolonged hospital stays [2], and significantly higher medical costs and resource utilization [3] [4].

The molecular epidemiology of MRSA has been extensively studied globally, utilizing various typing methods. These include SCCmec typing, which classifies isolates based on the essential components of the *mec* complex, *spa* typing, and sequence typing (ST) as defined by multilocus sequence typing (MLST). Clinical MRSA isolates from humans are predominantly categorized into SCCmec types I through V and assigned to clonal complexes (CCs), including CC1, CC5, CC8, CC22, CC30, among others [5]. Initially, MRSA was predominantly associated with healthcare settings, causing hospital-acquired MRSA (HA-MRSA) infections [6, 7]. However, the late 1990s marked a significant epidemiological shift with the emergence of community-acquired MRSA (CA-MRSA), with infections in healthy individuals with no prior healthcare exposure [8]. Molecular epidemiological studies have revealed distinct characteristics of HA-MRSA and CA-MRSA with differences in their SCC*mec* types and antibiotic susceptibility profiles. CA-MRSA is primarily associated with SCC*mec* types IV, V, or VII and tends to retain susceptibility to several non-β-lactam antibiotics. In contrast, HA-MRSA is characterized by multidrug resistance phenotype and typically harbors SCC*mec* types I, II, or III [9]. Over recent years, the prevalence of CA-MRSA clones has significantly increased, and in many regions, these CA-MRSA lineages have overtaken HA-MRSA as aetiological agents of nosocomial infections, thus highlighting their growing impact on public health.

The available data on the incidence, molecular characteristics, and mortality rates of MRSA in the Middle East highlight a significant burden in the region [10, 11]. Studies from countries such as Kuwait [12, 13], the United Arab Emirates (UAE) [14], Saudi Arabia[15, 16], and Qatar[17] reveal wide clonal diversity of MRSA. As a major member of the Gulf Cooperation Council (GCC), the Kingdom of Saudi Arabia (KSA) spans a vast geographical area and is characterized by a highly dynamic and diverse population, influenced by substantial immigration and religious tourism. A previous meta-analysis demonstrated a high prevalence of MRSA in KSA, particularly in the western region [18]. Molecular studies from KSA and neighboring GCC countries (Kuwait, UAE, Oman and Qatar) have identified a wide range of MRSA sequence types (STs). ST5, a globally significant lineage, is frequently reported in the region, often associated with *spa* types t688 and t002 and varying levels of *Panton-Valentine leukocidin* (*pvl*) gene positivity. ST8, commonly linked to the USA300 clone, has been detected in Saudi Arabia and adjacent countries, typically *pvl* gene -positive and associated with *spa* type t008. ST22, a predominant lineage in healthcare settings, is also widely reported in KSA, Kuwait and UAE, displaying considerable *spa* type diversity. Other lineages, including ST6, ST80, ST88, ST30, ST1, and ST97, have also been documented, exhibiting distinct genetic traits such as differences in *spa* types and PVL profiles. While these studies indicate the growing prevalence of CA-MRSA clones in KSA, they have primarily relied on traditional molecular techniques, which lack the resolution of whole-genome sequencing (WGS). This limitation hampers direct comparisons and accurate lineage tracking due to inconsistencies in reporting. Moreover, most studies have been restricted to individual hospitals or regions, leaving the national population diversity of MRSA largely unexplored. These gaps underscore the need for a comprehensive, WGS-based investigation to provide a more detailed and precise understanding of the molecular epidemiology of MRSA in KSA.

To address this gap, we conducted a nationwide genomic epidemiology study on a systematically collected, large-scale dataset of *S. aureus*, predominantly composed of MRSA strains, from 34 Ministry of Health (MOH) hospitals across Saudi Arabia. Leveraging whole-genome sequencing with both short- and long-read technologies, we achieved high-resolution characterization of MRSA population diversity and explored the genomic determinants of antimicrobial resistance and virulence. Our findings represent the first comprehensive genomic analysis of MRSA epidemiology in Saudi Arabia, revealing the expansion and ongoing evolution of CA-MRSA clones. These evolutionary processes are tightly linked to the concurrent acquisition of plasmids and key virulence genes, emphasizing the interplay between clonal adaptation and horizontal gene transfer.

## Methods

### Sampling and collection

This study received ethical approval from the Institutional Review Board (IRB) of King Abdullah University for Science and Technology (approval number 23IBEC027) and IRB of Saudi Ministry of Health (approval number: 23-23 M). As a component of a national surveillance initiative, the reference laboratory at the Saudi Ministry of Health collected samples from hospitals affiliated with the Ministry through standard hospital protocols between January 2022 and 2024. To ensure data integrity, isolates were deduplicated, with only one sample included per patient. A total of 36,286 isolates were identified as *S. aureus*, of which 98% were confirmed to be MRSA.

### Strain identification and susceptibility testing

Identification and antimicrobial susceptibility testing were conducted using the Vitek2^®^ system (bioMerieux, Marcy-l’Etoile, France) in accordance with manufacture provided protocol For susceptibility testing, the AST-GP67 cards with a panel of a range of antimicrobials was used. Among the β-lactams, the panel included cefoxitin (FOX), benzylpenicillin (PEN), ampicillin (AMP), amoxicillin/clavulanic acid (AMC), ampicillin/sulbactam (SAM), piperacillin (PIP), piperacillin/tazobactam (PTZ), and oxacillin (OXA). Cephalosporins tested were cefaclor (CEC), cefixime (CFM), cefotaxime (CTX), ceftazidime (CAZ), and cefepime (FEP). Carbapenems included imipenem (IMP) and meropenem (MEM). Aminoglycosides assessed were gentamicin (GEN) and streptomycin (STR). Fluoroquinolones included ciprofloxacin (CIP), levofloxacin (LVX), moxifloxacin (MXF), and ofloxacin (OFX). Macrolides and lincosamides were represented by azithromycin (AZM), clarithromycin (CLR), erythromycin (ERY), and clindamycin (CLI). Other tested antimicrobials included quinupristin/dalfopristin (QDA), linezolid (LNZ), vancomycin (VAN), tetracycline (TET), tigecycline (TGC), nitrofurantoin (NIT), rifampicin (RIF), and trimethoprim/sulfamethoxazole (TMP/SMX). Phenotypic resistance was defined according to the Clinical and Laboratory Standards Institute (CLSI) breakpoints based on minimum inhibitory concentration (MIC) values [20].

### Sequencing, assembly, mapping and GWAS analysis

Isolates were cultured overnight at 37°C in Luria–Bertani (LB) broth. Genomic DNA (gDNA) was extracted using the DNeasy Blood and Tissue Kit following the manufacturer’s protocol (QIAGEN, Hilden, Germany). The quality of the extracted gDNA was assessed using a DS-11 DNA spectrophotometer (Denovix, US), while its quantity was measured using a fluorometric method with a Qubit 4.0 fluorometer and a high-sensitivity double-stranded DNA assay kit (Thermo Fisher Scientific, US). Genomic libraries were prepared for 612 isolates using the MGIEasy Fast FS DNA Library Prep Kit, adhering to the manufacturer’s instructions (MGI Technology, China). Enzymatic fragmentation was employed during library preparation, followed by library denaturation and circularization. Whole-genome sequencing (WGS) was performed on the DNBSEQ-G400 platform (MGI Technology, China) using a 2×150 bp paired-end read protocol. To prepare sequenced libraries for long-read sequencing, we employed 96-plex Rapid Barcoding Kits for multiplexing. These libraries were then loaded into PromethION flow cells (Oxford Nanopore Technologies) and subjected to a 72-hour run following the manufacturer’s protocol.

The short reads underwent quality control using the FastQC package in R (v0.1.3). Genomes were assembled using the Unicycler *de novo* assembly pipeline (v0.5.0) (www.github.com/rrwick/Unicycler) with default settings [19]. Assemblies were processed, and contigs shorter than 200 bp were removed from the analysis. Genomes were profiled and characterized using Bactopia (v3.1.0) (www.bactopia.github.io) to determine the sequence types, SCCmec types, and spa types [20]. We also used AMRFinderPlus [21] pipeline, with identity coverage cut-off values of 50%, for identifying resistance genes.

We annotated the *de novo* assemblies using Prokka (v1.14.5) [8] and utilized Panaroo for pangenome reconstruction [9]. For the phylogenetic analysis, we aligned the short-read sequences to the reference genome of *S. aureus* NCTC 8325 (accession number: PRJNA57795), employing the Snippy pipeline (available at https://github.com/tseemann/snippy) with its default parameters. We calculated pairwise SNP distances from the core genome alignments. To assess genetic diversity within each province, we determined the average SNP distance by calculating the mean number of SNP differences between all pairs of isolates from the same province.

We contextualized our isolates using the Pathogen Detection database (https://www.ncbi.nlm.nih.gov/pathogens/). On 06/10/2024, we retrieved epidemiological SNP clusters from the database, which included genomes with pairwise SNP distances of up to 50 SNPs. The clustering was performed by the Pathogen Detection portal’s automated high-throughput pipeline (https://www.ncbi.nlm.nih.gov/pathogens/pathogens_help/#references).

We assessed the significance of associations between accessory genes, SNPs, and resistance phenotypes while accounting for population structure using Scoary (v1.6.16) [22]. This analysis was based on the Panaroo output for accessory genes and SNPs identified through post-read mapping to the reference genome. We specifically evaluated pairwise p-values (both worst and best) to be smaller than 0.05. These pavlaues are adjusted to account for the confounding effects of population structure, such as lineage effects.

### Transmission analysis and phylodynamic analysis

For the most prevalent *spa* types within the largest sequence type (ST) clones—ST5, ST8, ST80, ST88, and ST672—we performed phylodynamic analyses to estimate key epidemiological parameters for each clone. These clones were ST8-t008, ST88-t690, ST672-t3841, ST6-t304 and ST5-t311. To enhance our dataset and improve the temporal signal, we integrated our genomes with those sequenced in a single hospital study in Jeddah [23].

We selected the isolates with the best assembly metrics, specifically the highest N50, for each clone. The contigs of these selected strains were merged to create local reference genomes. Short reads from each strain within the clone were aligned to these reference genomes to generate a core genome SNP alignment. This alignment was processed using Gubbins (v3.3.1), with five iterations to remove hypervariable regions.

To infer the ancestral origins of the main clones within the SNP clusters, we conducted phylogeographic diffusion analysis in discrete space using BEAST (v2) [24]. The city of isolation served as the discrete state for each taxon, and we applied a constant population size model with uniform priors on the clock rate. Moreover, a symmetric model with a uniform prior distribution was used for the discrete trait substitution model to analyze the spatial diffusion of the clones. We evaluated the convergence of the Markov chain Monte Carlo (MCMC) chains by ensuring that the effective sample size (ESS) for critical parameters was greater than 150.

To investigate changes in population size over time, we applied a nonparametric growth model to sample population sizes along the dated phylogenetic tree for the five clones. This analysis was performed using the skygrowth.mcmc function, and the results were visualized with the plot function within the Skygrowth package (v0.3.1) [25]. For the ST8-t008 clone, we reported the age and population growth for the clade containing the *pvl* gene as well. For the ST6-t304, we exlcuded two divergent genomes.

To validate the results from the phylodynamic analysis, we reconstructed genealogical networks of the sampled sequences, assuming co-sampling of ancestors and descendants for the five clones. This was attained using the adegenet package in R (v1.3-1) [26], which optimizes the likelihood of the networks based on pairwise SNP distances from core genomes and the corresponding isolation dates. SNP distances were calculated from the BEAST input alignments, where hypervariable sites were excluded. These distance metrics, along with the collection dates, were input into the seqTrack function of the adegenet package. To define genetic relatedness, we applied a 22-SNP cut-off, a threshold previously demonstrated to capture 95% of epidemiologically linked cases within six months [27]. The resulting genealogical networks were visualized using the igraph library in R [28].

### Plasmidome analysis

We conducted third-generation sequencing on selected representative samples to characterize the antimicrobial resistance (AMR)-linked plasmids and improve typing within the collection. Forty isolates with the highest number of resistance genes were chosen, representing diverse clades and distinct resistance patterns. Moreover, long-read sequencing was performed on seven untypable isolates, and their profiles were submitted to the PubMLST database (www.pubmlst.org) to obtain new sequence type (ST) codes. The list of the isolates subjected to long-read sequencing is provided in Supplemental Table S1.

For library preparation, we utilized 96-plex Rapid Barcoding Kits for multiplexing, loading the libraries onto PromethION flow cells (Oxford Nanopore Technologies) for a 72-hour sequencing run, following the manufacturer’s instructions. Hybrid assemblies were generated using Unicycler with the conservative option. The resulting contigs were screened for full copies of origins of replication, virulence factor genes, and AMR genes using BLAST, referencing relevant databases. Visualization and validation of assembled genomes were performed using Bandage (v0.9.0) [28].

We extracted plasmid fragments containing resistance genes, which were then visualized and annotated using the built-in tools of the Proksee portal (www.proksee.ca) [29]. Resistance genes were identified through BLAST searches on the AMRFinderPlus [21] database. To find clusters within *blaZ*-containing plasmids, we conducted blast search and identified plasmids with identical plasmid replicons. To confirm the presence of these plasmids in other isolates not selected for long-read sequencing, we mapped the short reads of these strains against the extracted plasmid fragments, with mapping coverage exceeding 90% serving as confirmation. We visulized phylogenetic trees, along with antimicrobial resistance and virulence factor genes and plasmids with the ggtree package (v3.8.2) in R [29].

### Statistical significance tests

We conducted statistical significance tests using R. A one-way proportion test was employed to evaluate differences in ratios, while the one-way Wilcoxon signed-rank test was used to assess differences between means. For parameters inferred from the Bayesian analysis, significance was determined by examining the 95% credible intervals (highest posterior density [HPD]), representing the shortest interval encompassing 95% of the probability density. We analyzed the genetic diversity and incidence of MRSA in each province in relation to population size, using demographic data from www.citypopulation.de/en/saudiarabia/cities/, retrieved on 05/06/2024.

### Data availability

Genomic data collected in this study were deposited in the European Nucleotide Archive (ENA) under the study accession number PRJEB71150. The assemblies were uploaded to the NCBI GenBank database under the accession number PRJNA1050907. Detailed metadata associated with the genomes are available in Supplemental Table S1. All the intermediate files and codes are provided in the GitHub directory for the project: www.github.com/gzhoubioinf/MOH_MRSA.

## Results

### Overview of the MRSA and MSSA infection rates across the KSA

We conducted a genomic survey of 612 *S. aureus* isolates recovered from different body sites across 34 hospitals in a nationwide hospital network. The prevalence of *S. aureus* exhibited regional variability across provinces; however, no significant correlation was observed between incidence of *S. aureus* and the number of inhabitants of each province (p-value from Spearman’s rank correlation test > 0.05). Out of the 612 isolates, 30 lacked the *mecA* gene and were therefore classified as MSSA (methicillin-susceptible *S. aureus*). These figures made our collection representative of 1.5% and 4% of the total 35,560 MRSA and 726 MSSA isolates, respectively. The frequency of MSSA and MRSA was not significantly different in wound (16% MRSA versus 27% MSSA) and blood (66% MRSA versus 44% MSSA) (p-value > 0.01 from proportion test), showing that both strains could resides in similar sites. Isolates were selected to maximize geographical and temporal representation.

### High population diversity with dominant clones showing body site-specific preferences

The collection was diverse, comprising 48 distinct sequence types (STs). Of these, nine STs were novel, for which new ST identifiers were obtained (Suuplumental Table S1). Despite the high diversity, a few ST clones dominated the collection, including ST8 (n=110), ST5 (n=63), ST88 (n=56), ST6 (n=52), ST672 (n=47), ST30 (n=46), ST97 (n=35), ST22 (n=30), and ST152 (n=30), each representing at least 5% of the population (Figure 1A). Isolates from these clones collectively accounted for 75% of the population. MSSA isolates were found in ST30 (n=2), ST5 (n=1), ST672 (n=3), and ST8 (n=1), while the remaining isolates were distributed among other STs. Six STs consisted exclusively of MSSA isolates (ST2867, ST291, ST45, ST7565, ST15, and ST1290). All prevalent STs were isolated from both wound and blood samples, except for ST152, which was not found in blood (Figure 1B). Despite the broad distribution across body sites, some clones exhibited strong associations with specific infection sites: ST5, ST97, and ST8 were more frequently associated with blood, while ST6 and ST152 were more commonly associated with wounds. This suggests a possible preference for specific infection sites, reflecting differences in pathogenicity or ecological adaptation, likely driven by specific virulence genes (see Discussion).

**Figure 1.**
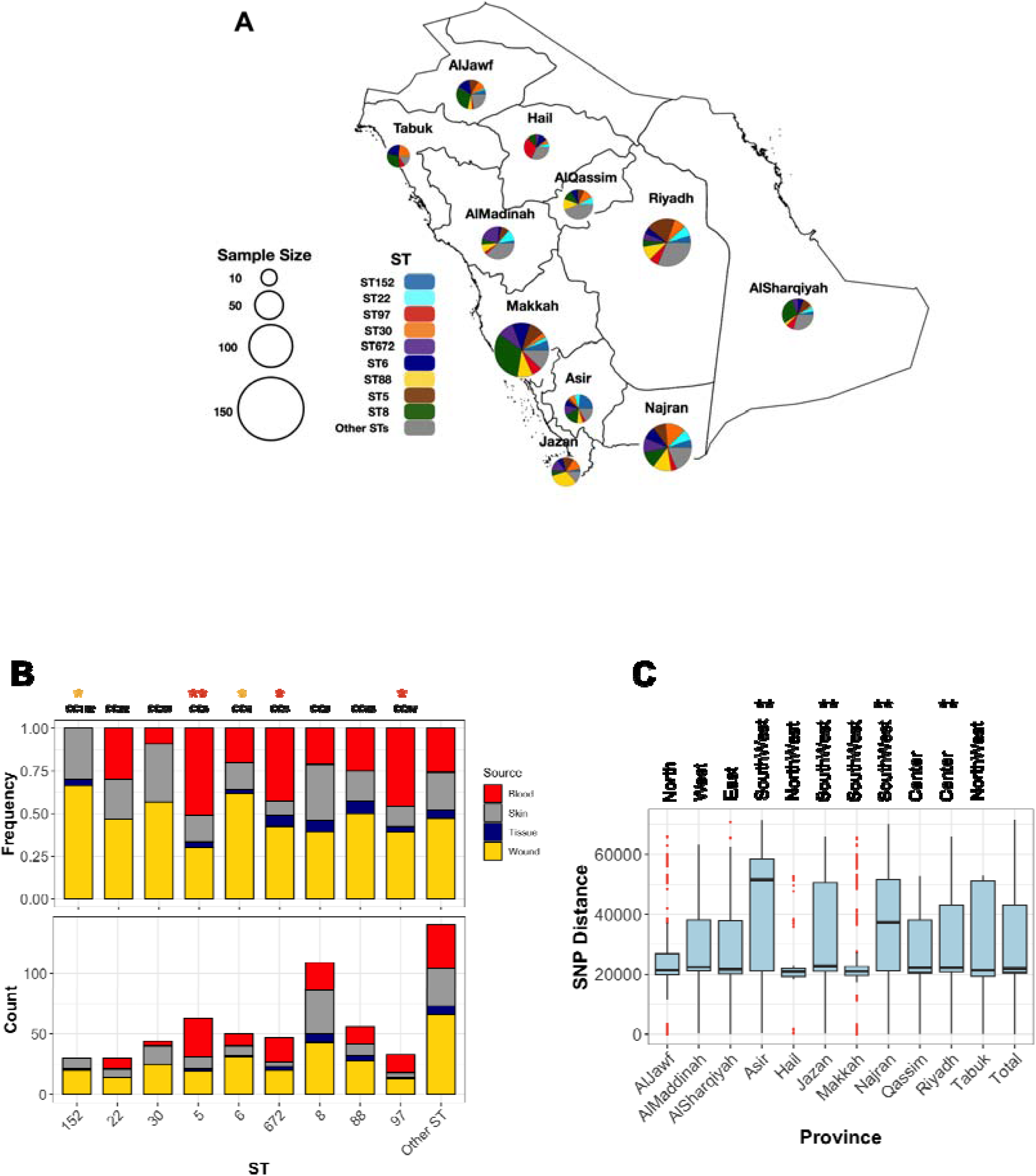
The distribution of major ST clones (STs with more than five representativ isolates) across sources and provinces. (A) Each pie chart represents the frequencies of major STs in the provinces of the KSA, with the chart size corresponding to the collection size from each province. (B) The relative (bottom) and absolute (top) frequencies of STs across body site of isolation for major STs. Asterisks (*) and (**) indicate significance levels of <0.05 and <0.01, respectively, based on a one-sided proportion test comparing the frequencies of blood (red) and wound (yellow) isolates. (C) Population diversity of the collection across provinces. Each boxplot represents pairwise SNP distances between genomes within a province. The ** symbol denotes a significance level of <0.01 based on a one-sided Wilcoxon rank test, comparing the SNP mean distances of each province to those of the other provinces.

### Widespread distribution and regional diversity of MRSA strains

The geographical distribution of different sequence types (STs) reveals that dominant clones are widely dispersed across the country. ST8 and ST6 isolates were identified in all provinces, while other clones—ST97, ST152, ST5, ST672, ST88, and ST97—were detected in most regions, specifically nine out of the eleven provinces (Figure 1A). Population diversity, based on SNP distance distribution, further highlights the broad genomic variation across provinces. The core genome population diversity, measured as the average pairwise SNP distances for isolates within the same province, exhibited a distribution comparable to the nationwide pattern (Figure 1C). Despite this overall similarity, regions in the central province of Riyadh and the southwestern part of the country displayed significantly higher diversity (p-value < 0.001, one-sided Wilcoxon rank test). These provinces, characterized by larger human populations and more diverse demographics, may facilitate the introduction and circulation of a wider variety of MRSA strains.

### Diverse Antimicrobial Resistance Profiles and Genetic Determinants Among MRSA Clones

While the collection was broadly resistant to β-lactams, variation in resistance levels was observed for other antimicrobial classes. ST8 and ST30 clones were significantly more resistant to macrolides (p-value from proportion test < 0.01). ST8, along with ST5 and ST772, showed higher resistance to ciprofloxacin (Figure S1). ST5 and ST22 isolates were more resistant to trimethoprim, while ST97 and ST152 were frequently resistant to aminoglycosides. Resistance to last-resort antimicrobials, vancomycin and linezolid, was observed in three ST8 and ST88 isolates, and in two isolates of ST5 and ST834, respectively. The resistome analysis further revealed a variable distribution of antimicrobial resistance determinants, with ST8, ST30, and ST152 harboring a greater number of resistance determinants compared to the rest of the population (p-value < 0.001, one-sided Wilcoxon rank test) (Figure 2A). Out of 47 antimicrobial resistance genes/mutations, 27 determinants were more frequent in at least one sequence type (ST) (Figure 2B). Several of these resistance genes/SNPs demonstrated strong associations with resistance phenotypes after adjusting for population structure. (pvalue from GWAS analysis < 0.01). These included the bifunctional *aac(6’)/aph(2’’)* aminoglycoside phosphotransferase, which confers resistance to aminoglycosides and was prevalent in ST97 and ST152; *msr(A)*, an Msr family ABC-F type ribosomal protection protein, and *ermC*, a macrolide-lincosamide-streptogramin B resistance protein, both of which were prevalent in ST5, ST8, and ST30 and confer resistance to macrolides; the trimethoprim-resistant dihydrofolate reductase *dfrG*, prevalent in ST5, which confers resistance to trimethoprim; and the putative tetracycline resistance pump *tetK*, underlying tetracycline resistance in ST88 and ST30 (Figure 2B). Genomic context analysis from long-read squencing data confirmed plasmid contexts for these genes, except for *dfrG*. At the SNP level, distinct evolutionary trajectories for ciprofloxacin resistance were observed across lineages. These included nonsynonymous mutations such as S80F in DNA topoisomerase IV subunit A (*parC*), prevalent in ST22, ST5, and ST672, and S84L in DNA gyrase subunit A (*gyrA*), also prevalent in ST22, ST5, and ST672 (Figure 2B). Linozolid resistant isolates haboured the *cfr* gene encodes a 23S rRNA methyltransferase. The gene was situated on a a conjugative 38Kb plasmid that also carried the efflux pump protein *fexA*, conferring resistance to florfenicol (Figure S2). The conjugative plasmid showed >90% sequence identity to the first report of the *cfr*-*fexA* plasmid (accession KC206006) recovered from clinical strains in the US [30]. Although no *van* genes were detected, we identified potential missense resistance mutations, which could contribute to intermediate levels of vancomycin resistance. This includes mutations in the *vraR* gene (R121I, D59E), which is part of the cell-wall stress VraSR two-component regulatory system, as well as in the peptidoglycan biosynthesis *murF* gene (S350G, T91M, T301I) [31, 32].

**Figure 2.**
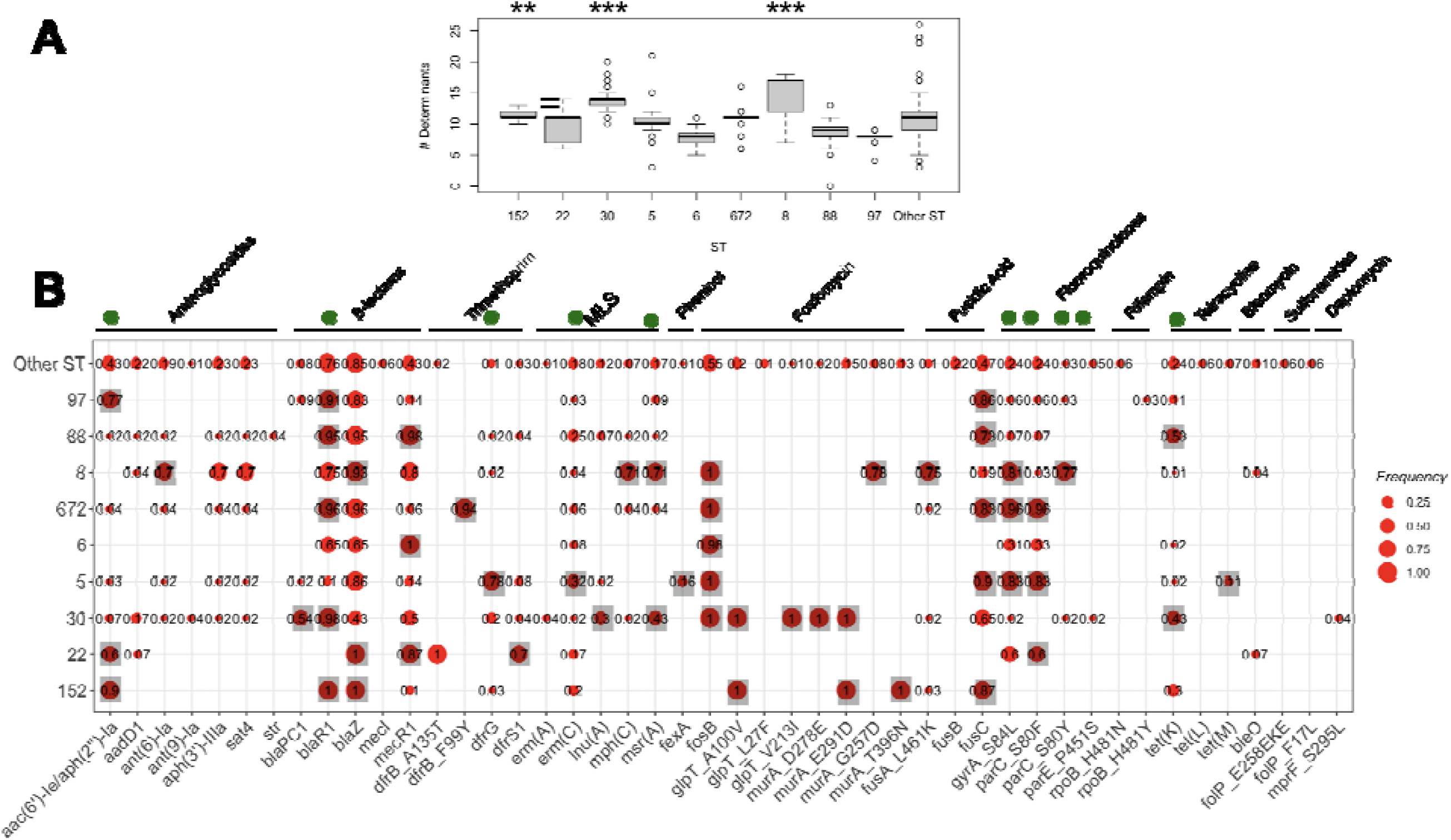
The frequency of A) resistance determimants counts and B) anticmicrobial resistance genes/mutations across the major STs. Each boxplot in A) denotes the count of resistance determinanats identified by AMRFinder. The ** and *** signs corresponds to the significance levels of <0.01 and <0.001, respectively, from the one-sided proportion test, indicating whether the mean of the frequency of resistance determinanats count were higher in the ST clone compared to the rest of the collection. The green circles on top genes in B) show the determinants signifintly linked with the resistance phenotype from the GWAS analysis. The grey squares in B) shows genes/mutations which had a siginficantly higher frequency in each ST compared to the rest of the collection (p-value<0.01 from one-sided proportion test).

### Distinctive virulence profiles distingusies CA-MRSA clones

The screening of virulence genes revealed a higher count in the ST8, ST5, and ST22 clones compared to the rest of the collection (Figure S3A). Of the 45 virulence factor genes identified, 34 showed variable presence across clones (p-value < 0.05, Fisher’s exact test) (Figure S3B). Distinct patterns emerged, encompassing toxins, adhesins, superantigens, and enzymes. The *cna* collagen-binding adhesin gene, which aids S. aureus in adhering to host tissues, was overrepresented in wound-associated ST152 and ST clones, although the role was also shown in bloodstream infections [33]. Twenty-four of the 29 superantigen genes, known for producing potent immunostimulatory exotoxins linked to bloodstream infections [34, 35], were variably present but overrepresented in at least one major clone compared to minor clones (Figure S3B). These superantigen genes contributed to the higher virulence gene count in bloodstream-associated ST5 and ST8 clones. The superantigen TSST-1, associated with toxic shock syndrome—a hallmark of CA-MRSA infections—was detected in 43 out of 612 isolates (7%) across eight sequence types, with significant prevalence in major ST22 strains and sporadic presence in ST5, ST672, and ST8 clones. Another hallmark of CA-MRSA infections, the pore-forming cytotoxin Panton-Valentine Leukocidin (PVL) genes (lukSF-PV), which target white blood cells, were present in 42% (258/612) of genomes across 21 of the 45 STs, including 4 novel STs we reported. This locus was found in all major clones except ST97. The majority of ST152, ST30, and some lineages of ST88, ST30, and ST22 carried these genes, although their frequency was lower in the ST5, ST672, and ST6 clones (Figure S3B). The differential distribution of virulence factor genes highlights the adaptation of specific MRSA clones to various infection types and host environments.

### Different types of mobile *mecA*-carrying chromosomal cassettes distinguish sublineages within each clone

Each clone in the population exhibited diverse *spa* and SCCmec types. The dominant *spa* types for ST152, ST22, ST30, ST5, ST6, ST672, ST8, ST88, and ST97 were t355, t005, t021, t311, t304, t3841, t008, t690, and t267, respectively (Figure 3). Except for ST97, the dominant *spa* subtypes in other clones contained genomes with the *pvl* locus. The diversity of SCCmec subtypes within each clone suggests pervasive dissemination, coexistence, and replacement of these subtypes. While some ST22 genomes carried SCCmec type I, commonly associated with hospital-acquired MRSA, the majority of SCCmec loci in the major clones were smaller, CA-MRSA-associated types IVa (n=228), V/VII (n=197), and VI (n=17). The prevalence of these CA-MRSA SCCmec types, which are smaller than HA-MRSA locis, suggest the success of these clones, as suggested before [36].

**Figure 3.**
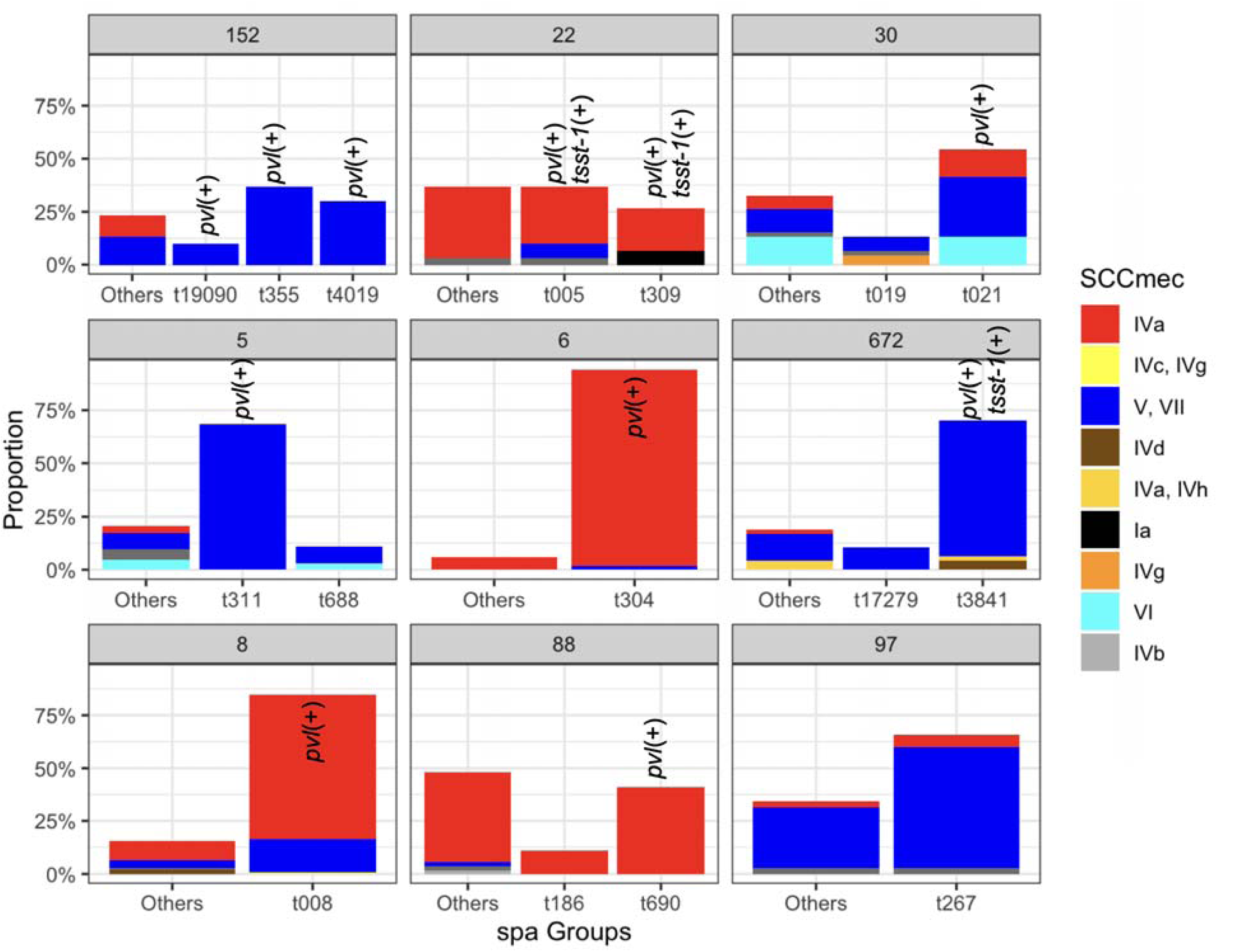
The relative frequencies of the spa types SCC*mec* types across the major STs. Spa types with a relative frequency of <0.1 for each clone were aggregated. Bars corresponding to the sub-clones which have at least one isolate carrying the *pvl* and *tsst-1* genes are marked.

### Temporal origins and population dynamics of the clones

After integrating *spa* and ST typing, we identified five major clones representing over 20% of the total collection. These included ST8-t008 (USA300 clone), ST88-t690, ST672-t3841, ST6-t304, and ST5-t311. We utilized the temporal signal within these clones to analyze their population dynamics. The results indicate a comparable molecular clock across the clones, ranging from 3.2 to 4.5 mutations per genome per year (Figure 4A). Despite the similar clock rates inferred from overlapping credible intervals, some clones, such as ST5-t311 and ST672-t3841, appear to have emerged more recently, with an estimated most recent common ancestor (MRCA) age of 20 to 30 years. ST8-t008 (USA300) clones were found to consist of two subclones, distinguished by the presence of *pvl* and resistance genes (Figure 5A). While the ST8-t008 lineage is long-standing (76 years), the *pvl*-positive subclade (ST8-t008*pvl+*) emerged relatively recently, within the past 15 years, making it younger than the other clones (Figures 4A and 5A). All clones exhibited an overall increasing population trend over time, although a slight recent decline was observed (Figure 4B) (see Discussion). ST5 and ST6 had larger population sizes; however, ST5, along with ST8 after the emergence of the *pvl+* subclone, showed the most rapid population expansion, increasing 1,000-fold over ten years (Figure 4B). Similarly, the ST88-t690 clone experienced a rapid population increase 12 years ago, coinciding with the emergence of a *fusC*-positive clone (Figure 4B, 5B). Overall, the findings indicate comparable molecular clock rates, effective population sizes, and the recent emergence of the five clones, which could be linked to the acquisition of resistance/virulence determinants, highlighting them as high-fitness, emerging clones.

**Figure 4.**
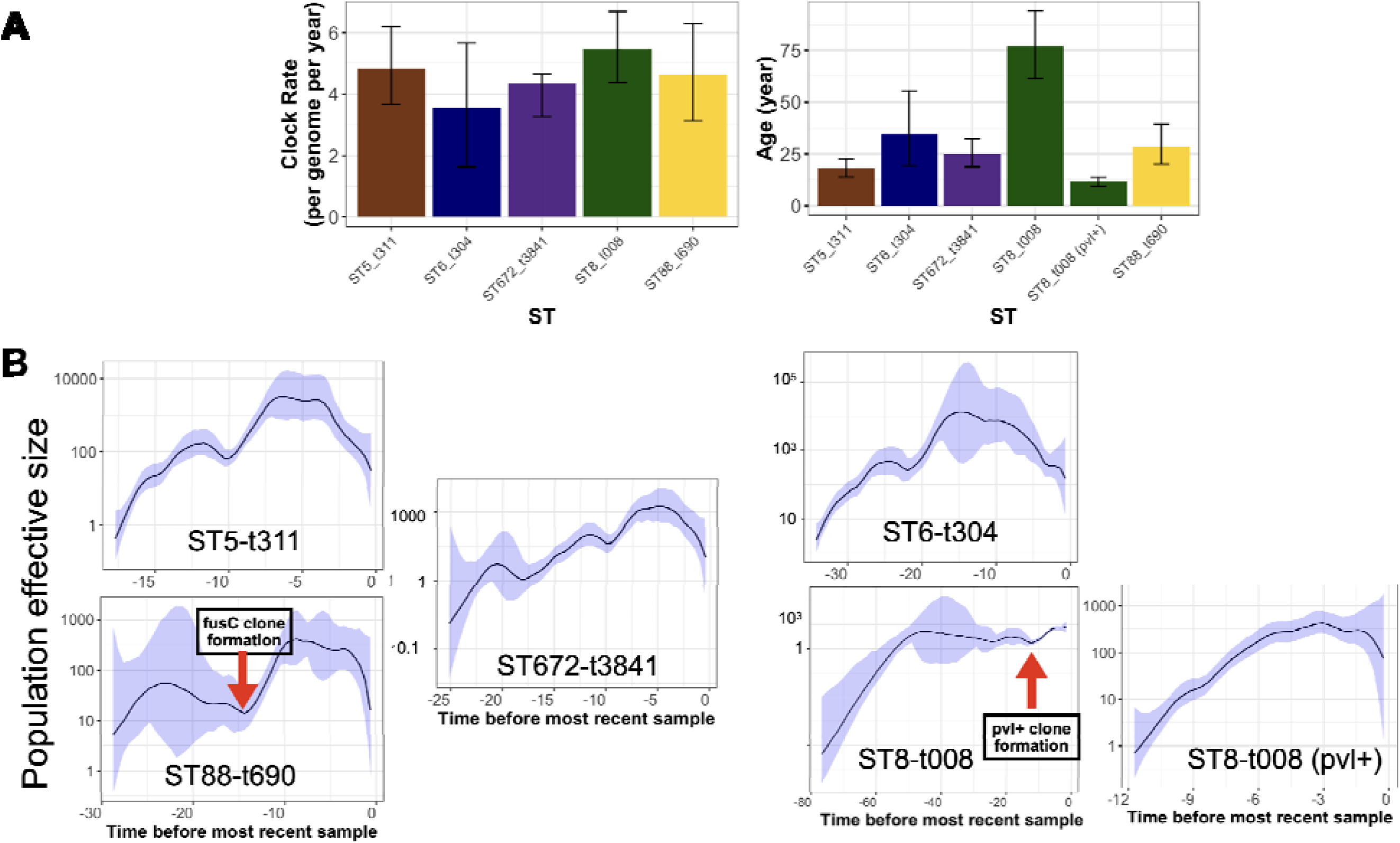
Phylodynamic analysis of the most prevalent spa type in the ST clones. A) The estimated clock rate and age of the clones from Bayesian analysis. The error bars are the credible intervals (95% Highest Posterior Density (HPD)). B) The skyline growth for the clones in A), based on the Bayesian tree for the clones. The shaded area corresponds to 95% confidence interval.

**Figure 5.**
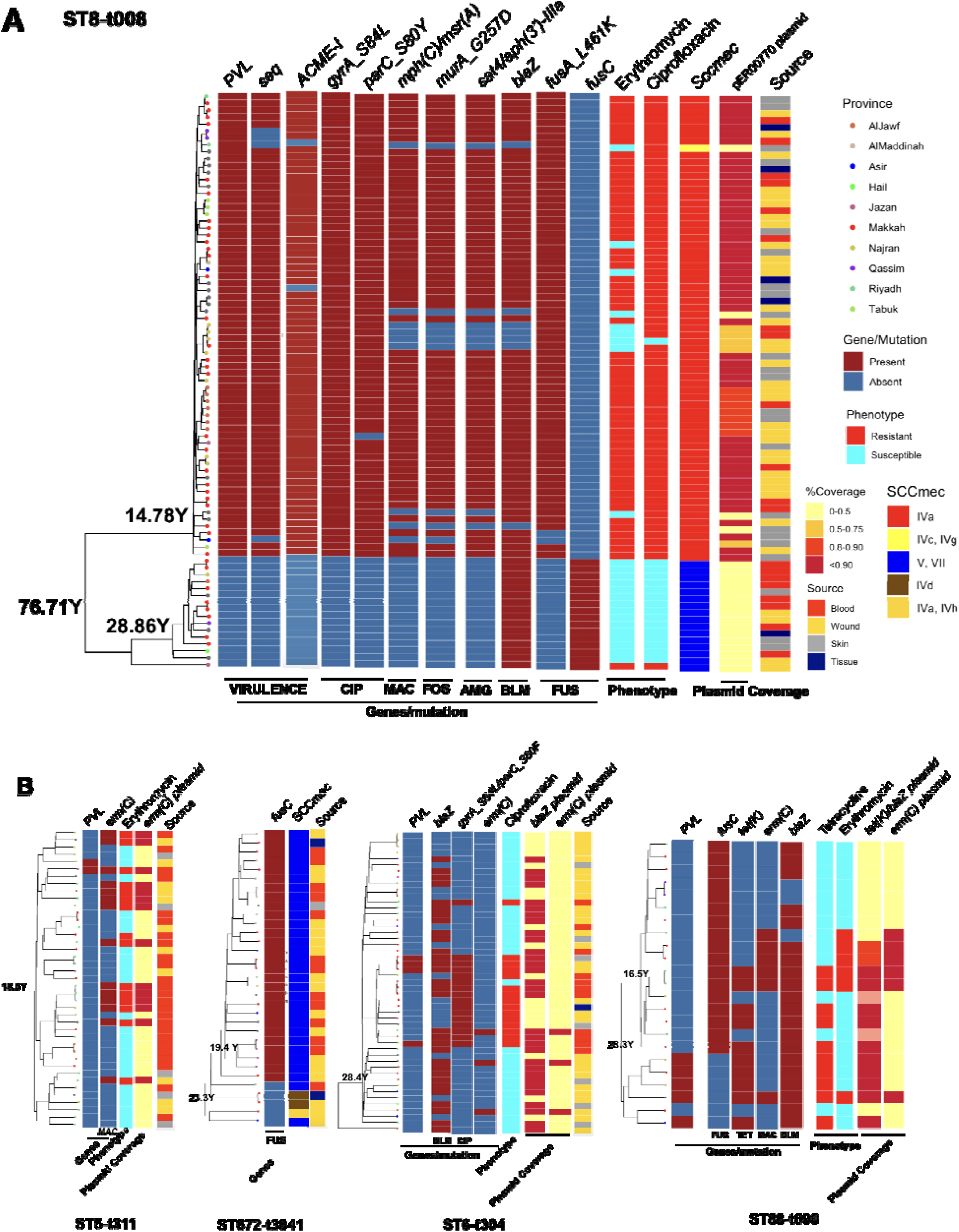
The phylogenetic trees for the clones with key resistance determinants, virulence factor genes, SCC*mec* type, antimicrobial resistance phenotype and the presence of key plasmids for the ST8-t008 clones in A) and other expanding clones B). We show resistance determinants and virulence factors and antimicrobial resistance phenotype that were variably present in each clone accroding to Figure 2, S1 and S3. The plasmid band shows the coverage percentage after mapping the short reads against the plasmid with the resistance genes retrieved from long-read sequencing data. Color codes in B) are the same as in A). The genomic map of the plasmids is provided in Figure S2.

### Expanding clones revealed the emergence of new clones linked with the acquisition of antimicrobial resistance plasmids

We next analyzed the dynamics of the expanding clones and integrated this with resistance determinants and plasmids identified through third-generation sequencing. In the USA300 clone, the subclone carrying *pvl* exhibited a significantly different composition of antimicrobial resistance genes (Figure 5A). While isolates from both subclones were recovered from different body sites, the larger *pvl*-positive subclone harbored SCC*mec* type IVa, whereas the *pvl*-negative subclone carried type VI, which was linked to the *fusC* gene (Figure 5A). The larger subclone also contained other key virulence genes (e.g., the superantigen *seq* and ACME-I gene) and resistance determinants (e.g., mutations conferring resistance to ciprofloxacin, fusidic acid, and fosfomycin, as well as genes for aminoglycoside and macrolide resistance). This resulted in increased resistance to macrolides (erythromycin) and ciprofloxacin (Figure 5A). The macrolide and aminoglycoside resistance genes in the *pvl+* clones were located on a 27kb *blaZ*-containing plasmid (identical to the clinical pER00770_1 plasmid) (Figure S2). Similar to the USA300 clone, other STs also exhibited mixing of isolates from different body sources, SCCmec replacement, the rise of clades with the *pvl* locus, and evidence of plasmid-borne resistance acquisition during clonal expansion (Figure 5B). The rise of *pvl(+)* lineages was observed in ST88-t690 and ST6-t304. Similar to the ST8 clade, ST672 contained two SCCmec types, with type V harboring the *fusC* gene, which emerged and expanded from a lineage with SCCmec IVd. The ST6-t304 and ST88-t690 clades acquired *blaZ*-containing plasmids with similar replicons, with the ST88-t690 clone also carrying the *tet(K)* gene (Figure 5B, S2). The acquisition of these plasmids was linked to phenotypic resistance to macrolides and tetracycline. Moreover, our results show the sharing of a 2.6kb *emr(C)*-containing plasmid (Figure S2) among subclades within ST5-t311, ST6-t304, and ST88-t690 clones (Figure 5B). These findings highlight the concurrent processes of clonal expansion and horizontal gene transfer, driving the enhancement of resistance and virulence in the expanding clones.

### CA-MRSA clones spread across the hospitals within the country

We then examined the spread of the five clones across hospitals by reconstructing the transmission network for each clone, assuming that ancestor and descendant genomes were sampled concurrently. Using a 20 SNP cutoff, which corresponds to hospital transmission over a six-month period (see Methods), we identified 93 transmission networks for the five abovementioned expanding clones, accounting for 15%-28% of the isolates within each clone (Figure S4). The largest proportion of isolates involved in a transmission network was observed for the ST8-t008 clone, which, except for the ST88-t690 clone, was higher than the other clones. The transmission links spanned all regions of the country for all clones (Figure S4). In addition to Saudi hospitals, ST8-t008, ST672-t3841, and ST88-t690 also showed recent mixing with European clinical samples, with three SNP clusters. ST88-t690 was mixed with Danish clinical isolates, with the closest SNP distance of 37 (SNP cluster PDS000144949.9), which, although not within the transmission cutoff, still suggests dispersion beyond Saudi Arabia. ST672-t3841 genomes were also clustered with Dutch clinical samples, with an SNP distance of 11 (SNP cluster PDS000170231.2), indicating that dispersion of the clone extends beyond KSA.

### Plasmidome analysis indicates diversity and horizontal transfer of *blaZ* containing plasmids

Our analysis of *blaZ*-containing plasmids, retrieved from long-read sequencing data, revealed significant diversity, which could be grouped into ten plasmid clusters based on their origins of replication (Table 1) (see Methods). The large diversity of replicons is also evident in the wide variation in plasmid lengths, which ranged from 12 Kb to over 42 Kb, with larger plasmids generally carrying a greater number of resistance genes (Table 1). These resistance plasmids exhibited various combinations of resistance genes significantly associated with resistance phenotypes, including *blaZ, blaI, blaR1, cadD, tet(K), and aac(6’)-Ie/aph(2’’)-Ia*, leading to the formation of multidrug-resistant plasmids. Except for one plasmid cluster, all clusters contained *cadCD* heavy metal resistance genes. The plasmids were distributed across different cities and body sites within five clusters. All plasmid clusters harbored OriT regions, as demonstrated for the blaZ-containing plasmids in the major clones in Figure S2, which enables plasmid transfers [37, 38]. Furthermore, a similar collection of plasmids was observed for five plasmid clusters of different ST types, suggesting the dissemination and sharing of plasmids across diverse genetic backgrounds (Table 1). The widespread distribution of plasmids across bacterial hosts and locations points to extensive plasmid transmission within hospital settings, likely facilitated by human carriers or hospital enviornments [39, 40].

**Table 1.**
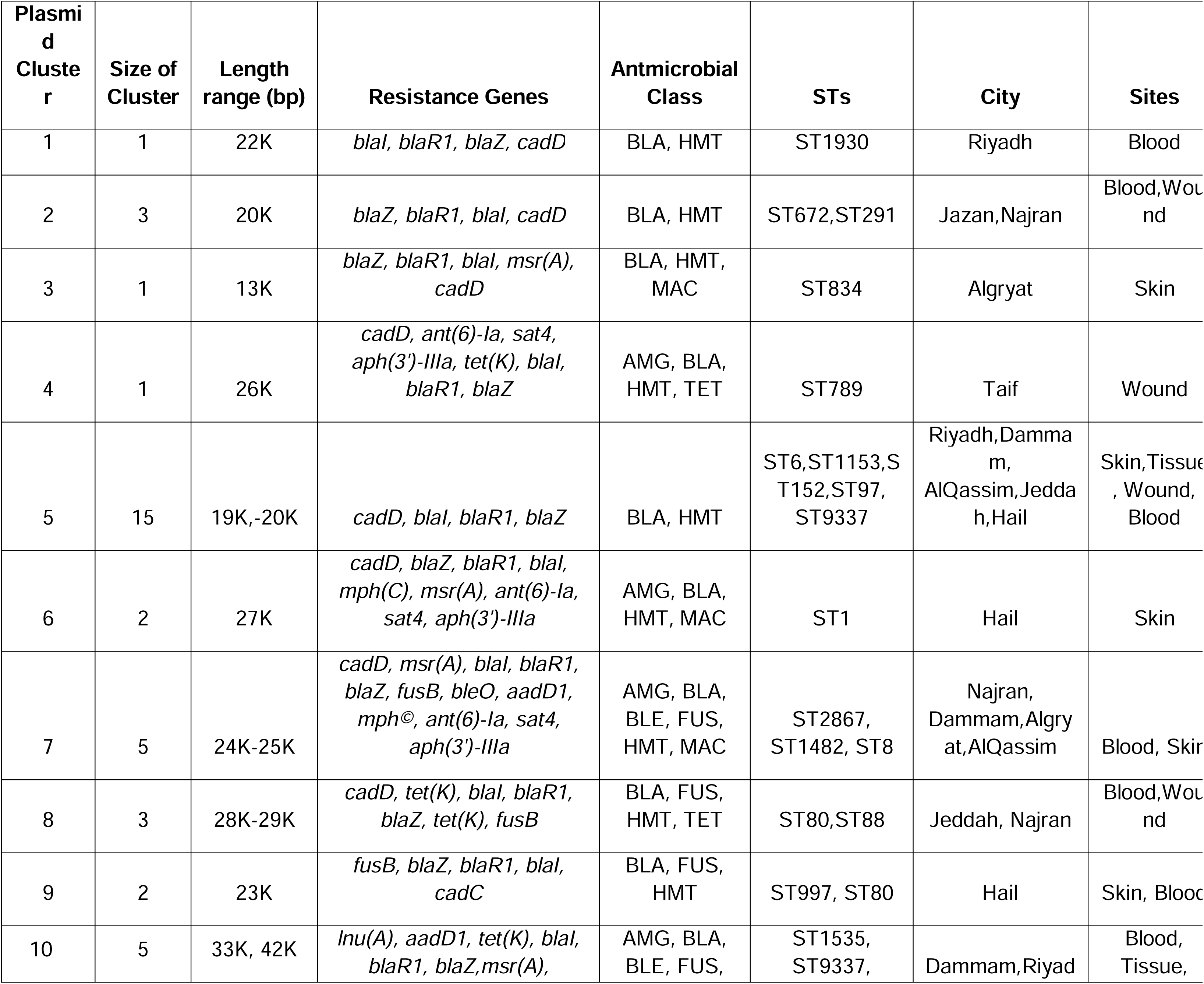
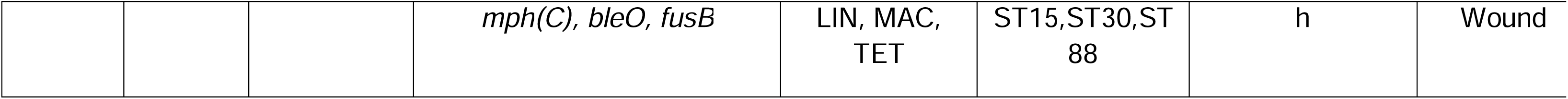
List of *blaZ*-containing plasmids retrieved from long-read sequencing data. Each group corresponds to plasmids with a unique origin of replication. The order of antimicrobial classes corresponds to the order of genes in the resistance genes column. The annotations for the abbreviations are as follows: BLA: β-lactam antimicrobials, HMT: Heavy metal tolerance, MAC: Macrolides, AMG: Aminoglycosides, TET: Tetracyclines, BLE: Bleomycin resistance, FUS: Fusidic acid, LIN: Lincosamides. The size of the cluster indicates the number of plasmids in the cluster.

## Discussion

### Summary

We conducted a genomic epidemiology analysis of a *S. aureus* collection, mostly comprised MRSA strains, retrieved from a large hospital network across the Kingdom of Saudi Arabia. Utilizing a range of genomic epidemiology techniques, we dissected the population diversity, identified novel MRSA strains, and investigated the recent evolution of emerging and expanding clones in the hospitals. By employing a hybrid sequencing approach, we uncovered extensive diversity and sharing of antimicrobial resistance (AMR)-linked plasmids between clones. We also revealed significant within-clone diversity and recent evolutionary events, including the concurrent acquisition of plasmid and clonal expansion. Given the diversity and breadth of the hospitals included in this study, we expect the findings to be generalizable to the broader Middle East and West Asia region.

### Population diversity of MRSA

The study revealed a genetically diverse population of co-circulating MRSA lineages in Saudi Arabia, carrying a wide array of genetic determinants linked to antimicrobial resistance and virulence. The dominant clones identified were either pandemic or regionally significant. ST8-t008 (USA300), the predominant clone, is a well-known pandemic CA-MRSA strain, particularly prevalent in North America but also reported in other regions, including the Middle East [13]. Similarly, ST6-t304, part of the CC6 CA-MRSA lineage, has demonstrated the ability to spread internationally, especially in Northern Europe [31, 41]. Other major clones were reported in Africa, East Asia, and the Middle East: ST88-t690 belongs to the ST88-IV African CA-MRSA clone [41], while ST672-t3841 is part of the CC361 lineage, and ST5-t311 is part of the CC5 clone, prevalent in the Middle East and South and East Asia [42–44]. These clones are associated with infections across various body sites and both hospital and community settings. The high level of genetic diversity of the MRSA population in Saudi Arabia likely reflects the country’s complex demographic landscape, shaped by its role as a major global travel hub and its highly diverse population, including over 30% expatriates. This contributes to the introduction and circulation of globally prevalent MRSA clones within the country.

### USA300 clone in Saudi shows similarity ot the prevalnt clone in the Americas

The ST8-t008 clone in our collection is predominantly composed of the USA300 variant, displaying all the hallmark characteristics observed in U.S. hospitals, including SCCmec type IV, the *Staphylococcus aureus* pathogenicity island 5 (SaPI5), and a multidrug-resistant 27 Kb plasmid [45, 46]. The formation dates for both the ST8-t008 and USA300 clones align with the reported ages of these lineages in the Western Hemisphere [47–49], suggesting that the expanding clone in Saudi Arabia mirrors the global expansion of the ST8 lineage, particularly the USA300 strains. The minor pvl(-) subclone of ST8-t008 with SCCmec V in our collection is less documented and has also been reported as a rare strain in a large-scale, single-hospital study in the U.S. [49]. Such spread and replacement of accessory resistance and virulence genes, including SCCmec subtypes were reported during hospital outbreaks and inter-hospital transmissions [50]. This is likely driven by the higher fitness of smaller SCCmec subtypes, along with the coselection and mobility of resistance genes [36, 48]. The coexistence of ST8-t008 subclones in our population may represent a snapshot of the population dynamics, where a fitter clone/subclone dominates and outcompetes others. Similar to previous studies, we observed a recent decline in population size of the clones, which has been previously attributed to improved infection control efforts in hospitals or the emergence of newly successful but rare clones [51]. To determine which factor underlies this dynamic pattern, a long-term longitudinal study incorporating epidemiological data, genomic surveillance, and transmission modeling would be necessary.

### Limitation and the need for One Health framework

Despite the breadth of our study, there are several limitations. Firstly, we lacked detailed clinical data, such as patient infection sources, travel history, infection types, and clinical outcomes. This limitation restricted our ability to draw conclusions about the clinical significance of emerging clones. Human mobility, particularly international travel , as it is common in KSA, plays a crucial role in the introduction and dissemination of both CA-MRSA strains, facilitating the spread of diverse clones within communities and healthcare settings and contributing to the importation and intermixing of MRSA reservoirs [50, 51]. Including information such as travel history in the analysis could help decipher the factors contributing to the spread of the clones. Secondly, the absence of environmental samples in our study did not allow us to determine the role of non-human reservoirs and agents in disseminating the strains across hospitals. Several common STs in our collection, such as ST97 (CC97)—primarily linked to livestock—along with ST6-t304, ST8-t008, and ST88, have been previously reported in animal sources globally, as well as in Saudi Arabia [41, 43, 52, 53]. A previous One Health study in Saudi Arabia reported the sharing of clones ST97, ST672-t3841, and ST5-t311 within a small genomic collection [43]. This potential overlap underscores the need for large-scale, systematic, and unified studies that integrate human, animal, and environmental reservoirs. Such studies are essential to uncover the relatedness and contributions of non-human sources to MRSA transmission at a national level.

### Conclusion

Our study underscores the importance of understanding the regional epidemiological characteristics of MRSA clones in a highly geographical setting, which remains significantly underrepresented in global pathogen genomic databases. Despite the limitations, we believe the current dataset provides a crucial baseline census of the circulating genetic diversity and antimicrobial resistance profiles of *S. aureus* in the Middle East. This contribution not only enhances our understanding of the local MRSA landscape but also aids in deciphering the global dissemination of high-risk and emerging MRSA clones., offering insights for international surveillance and control efforts.

## Acknowledgements

Acknolwedgment

G.Z., H.H., J.H., O.F., R.H., S.I., M.M., and D.M. were supported by KAUST faculty baseline fund (BAS/1/1108-01-01). AP is supported by KAUST baseline (BAS/1/1020-01-01). G.Z., H.H., J.H., O.F., R.H., S.I., M.M., and D.M. were also supported by FCC/1/5932-01-03 from KAUST Center of Excellence for Smart Health. The authors extend their appreciation to the Deputyship for Research and Innovation, “Ministry of Education” in Saudi Arabia for funding this research (IFKSUOR3-478).

## Authors Contribution

A.Y.A., G.Z., and H.H. conducted the research. D.M. and W.A.S. conceived the research and supervised the study. D.M., H.H, G.Z. and A.S. wrote the manuscript and interpreted the results. A.Y.A., J.H., O.F., R.H., S.I., A.A., M.M., M.B., A.A.A., A.A.R., M.A.A., S.S.A., D.B., A.N.A., A.T.A., Z.A.A., S.M.A., R.H.A., M.A., M.M.A., A.P., and A.S. contributed to research material and computational procedures.

## Decleration of conflict of interest

Authors declare no conflict of interest.

