## Supplemental Figures for "Methicillin-resistant *Staphylococcus aureus* in Saudi Arabia: genomic evidence of recent clonal expansion and plasmid-driven resistance dissemination"

### Supplementary Figures

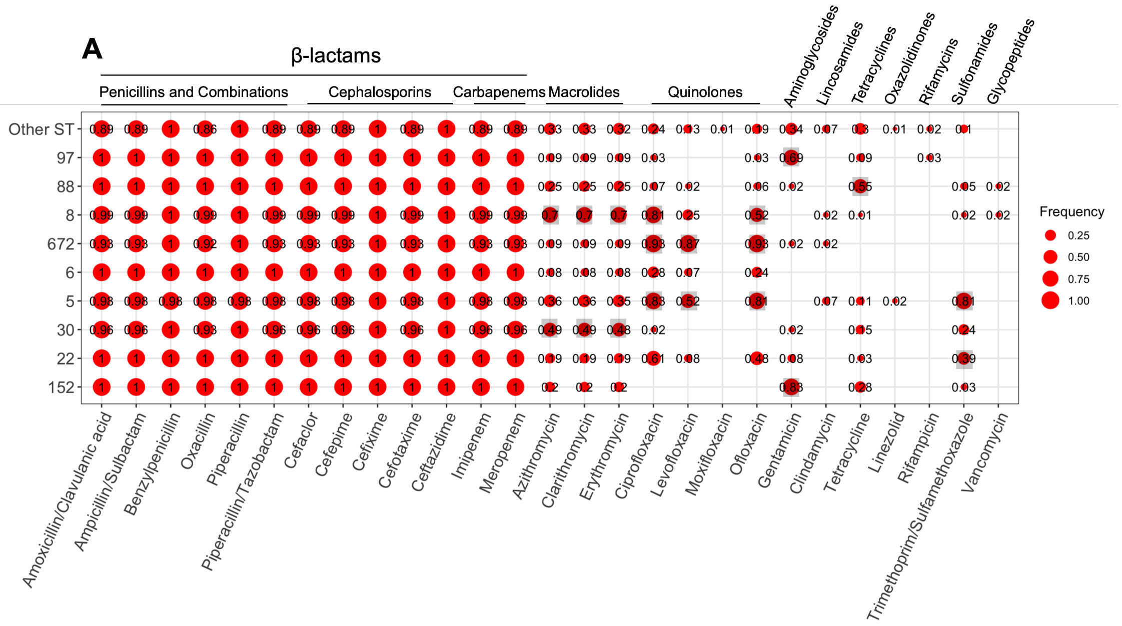


**Figure S1: The frequency of resistance phenotypes across major STs.** The grey squares indicate resistance levels that were significantly higher in each ST compared to the rest of the collection (p-value < 0.01 from a one-sided proportion test).

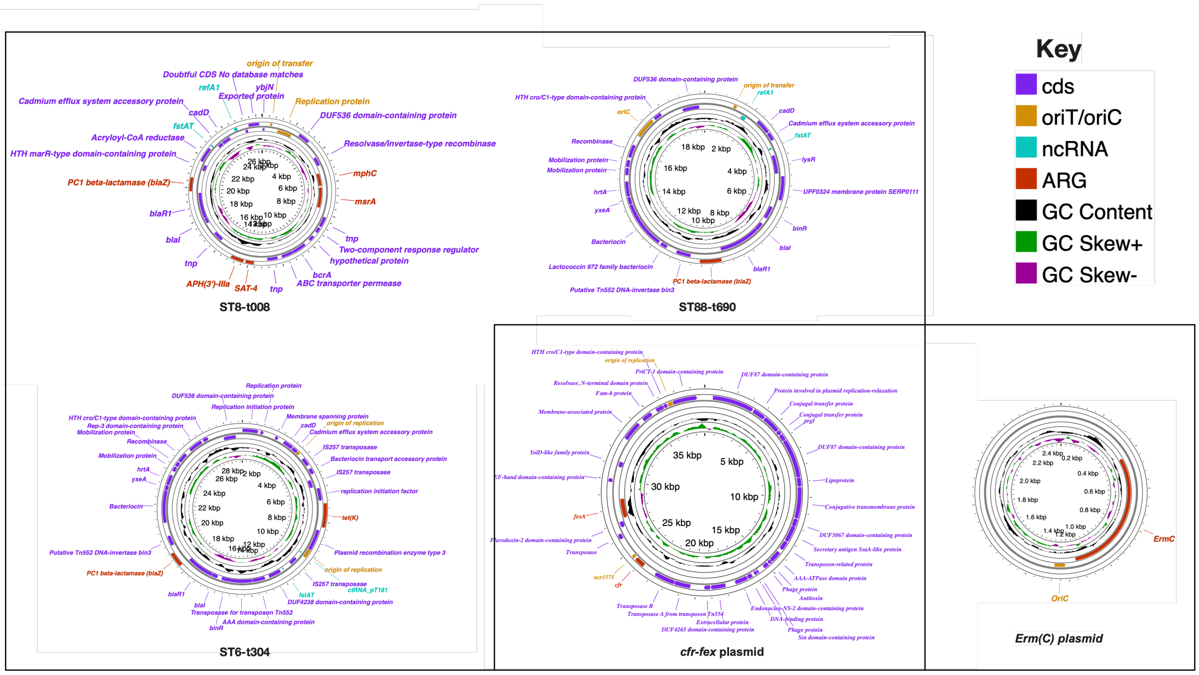


**Figure S2 The genomic map of the plasmids shown in Figure 6 and mntioned throughout the text.** The antimicrobial resistance genes (ARGs) were reported according to CARD. Figures were generated by www.proksee.ca built-in tools including annotation and ARG finding pipelines.

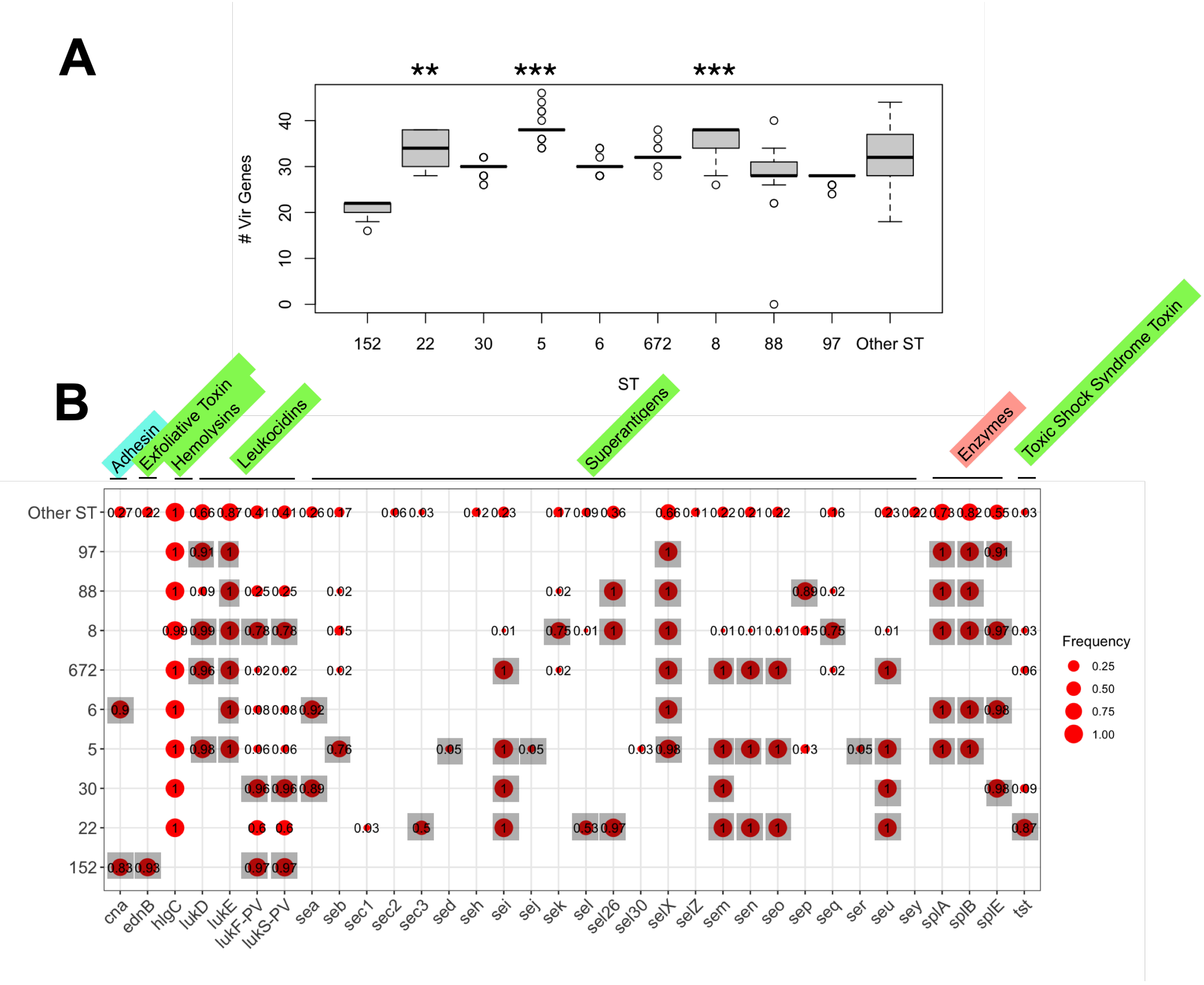


**Figure S3** A) The distribution of virulence factors across the major ST clones. Each boxplot denotes the count of resistance determinanats or viruelnce factor genes idenfitied srst2 tools (using VFDB), respectively. The ** and *** signs corresponds to the significance levels of <0.01 and <0.001, respectively, from the one-sided proportion test, indicating whether the mean frequsny of viruelnce factor genes count were higher in the ST clone compared to the rest of the collection. B) The frequency of virulence factor genes across the major STs. The grey squares shows genes/mutations/drugs which had a siginficantly higher frequency in each ST compared to the rest of the collection (p-value<0.01 from one-sided proportion test).

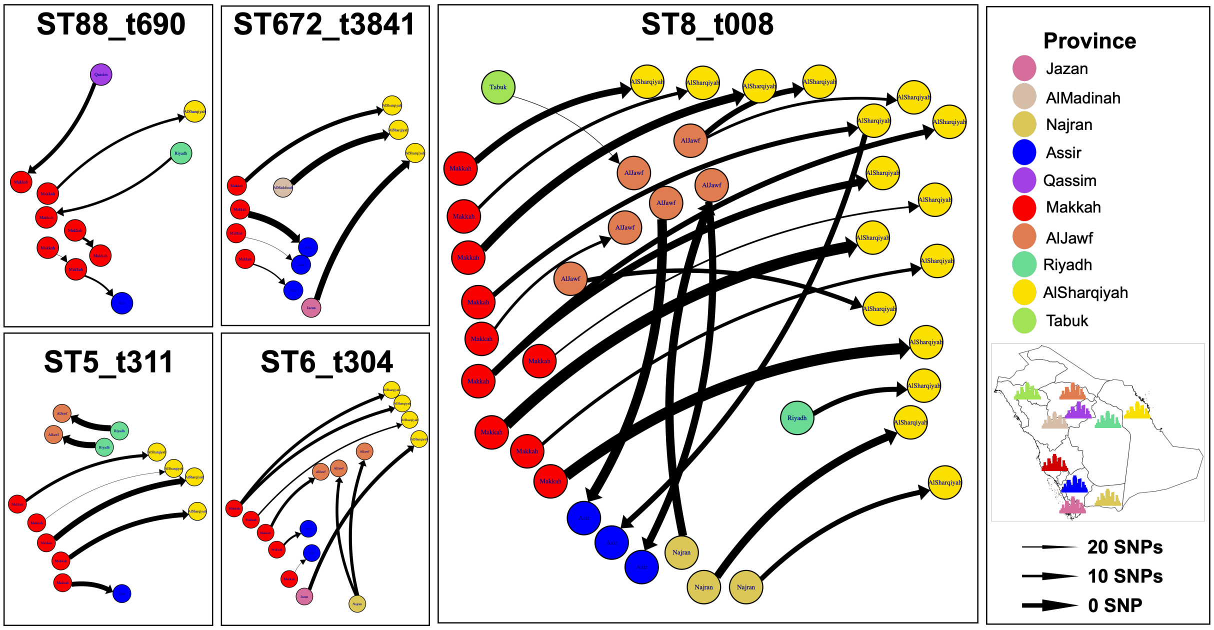


**Figure S4** The transmission network analysis for the identfied recent clones. The nodes and edges denote the isolates and transmission network, respectively. Colors illustrate provinces. The thickness of the edges denote the transmission link.
